# Endogenous oxytocin levels in children with autism: Associations with cortisol levels and oxytocin receptor gene methylation

**DOI:** 10.1101/2022.12.15.22283521

**Authors:** Evenepoel Margaux, Moerkerke Matthijs, Daniels Nicky, Chubar Viktoria, Claes Stephan, Turner Jonathan, Vanaudenaerde Bart, Willems Lynn, Verhaeghe Johan, Prinsen Jellina, Steyaert Jean, Boets Bart, Alaerts Kaat

**Affiliations:** KU Leuven, Department of Rehabilitation Sciences, Research Group for Neurorehabilitation, Leuven, Belgium; KU Leuven, Department of Neurosciences, Center for Developmental Psychiatry, Leuven, Belgium; University Psychiatric Centre, KU Leuven, Leuven, Belgium; KU Leuven, Department of Chronic Illness and Metabolism, Laboratory of Respiratory Diseases and Thoracic Surgery, Leuven, Belgium; KU Leuven, Department of Development and Regeneration, Research Group Woman and Child, Leuven, Belgium; KU Leuven, Leuven Autism Research (LAuRes), Leuven, Belgium; Luxembourg institute of health, Department of Immunology, Laboratoire National de Santé, Luxembourg

## Abstract

**Background:** Alterations in the brain’s oxytocinergic system have been suggested to play an important role in the pathophysiology of autism spectrum disorder (ASD), but insights from pediatric populations are sparse.

**Methods:** We examined salivary oxytocin in school-aged children with (n=80) and without (n=40) ASD (boys/girls 4/1), as well as characterizations of DNA methylation (DNAm) of the oxytocin receptor gene (*OXTR*). Cortisol levels were also assessed to examine links between the oxytocinergic system and hypothalamic-pituitary-adrenal (HPA) axis signaling.

**Results:** Children with ASD displayed altered (diminished) oxytocin levels in the morning, but not in the afternoon, after a mildly stress-inducing social interaction session. Notably, in the control group, higher oxytocin levels were predictive of lower stress-induced cortisol, likely reflective of a *<u>protective</u>* stress-regulatory mechanism for buffering HPA stress activity. In children with ASD, on the other hand, a more *<u>reactive</u>* stress regulatory mechanism was evident, involving a significant rise in oxytocin levels from the morning to the afternoon upon stress-induced cortisol release, i.e., to reactively cope with heightened HPA activity.

Regarding epigenetic modifications, no overall pattern of *OXTR* hypo- or hypermethylation was evident in ASD. In control children, a notable association between *OXTR* methylation and levels of cortisol was evident, likely indicative of a compensatory downregulation of *OXTR* methylation (higher oxytocin receptor expression) in children with heightened HPA axis activity.

**Conclusion:** Together, these observations bear important insights into altered oxytocinergic signaling in ASD, which may aid in establishing relevant biomarkers for diagnostic and/or treatment evaluation purposes targeting the oxytocinergic system in ASD.

## Introduction

Autism spectrum disorder (ASD) is a neurodevelopmental condition characterized by difficulties with social communication and interaction, combined with expressions of restricted and repetitive behaviors and interests. Aside these core ASD symptoms, individuals with ASD often also display a broad range of comorbid symptoms including attachment difficulties ^1^ and (social) stress and anxiety ^2^.

While the etiology and underlying neurobiological mechanisms responsible for the heterogeneous clinical presentation of ASD are not yet fully understood, recent lines of evidence suggest that alterations in the brain’s oxytocinergic system play an important role in the pathophysiology of ASD ^3^. Oxytocin is a nonapeptide produced in the paraventricular and supraoptic nuclei of the hypothalamus ^4^, from which oxytocinergic neurons project to various regions of the brain’s reward system and social brain circuits (e.g. amygdala) ^5^. Through its widespread neuromodulatory effects, oxytocin is identified as a key mediator of affiliative and prosocial behaviors, including interpersonal bonding, attachment and social attunement ^6–8^.

Initial preclinical studies have demonstrated links between aberrant oxytocinergic signaling and ASD symptom expression in rodent models of autism ^9–11^. However, investigating oxytocin concentrations in biological samples (mostly plasma and saliva) collected from humans with ASD, yielded a more mixed pattern of results (see meta-analysis of Moerkerke et al. (2021) ^12^). In particular, only young children with ASD (6-to-9 year old) showed reduced oxytocin levels compared to controls in four out of five studies ^13–17^, whereas none of the studies including 9-to-12 year old children with ASD reported a significant group difference in endogenous oxytocin levels ^18–20^. Moreover, studies including adolescents (11-to-16 year old) ^18–20^ and adults ^21,22^ with ASD revealed a mixed pattern of results, demonstrating no clear evidence for alterations in oxytocin levels in ASD.

Aside assessments of circulating oxytocin, also variations in epigenetic modifications of the oxytocin receptor gene (*OXTR*) have been examined in relation to ASD. The functioning of the circulating oxytocin is dependent on the functioning of the oxytocin receptor ^23^, which itself is encoded by the *OXTR* ^24^. DNA methylation (DNAm) is one of the most extensively studied epigenetic mechanisms involved in the regulation of gene transcription, such that increased DNAm frequency is generally associated with decreased gene transcription, and therefore less expression of the respective protein ^25^. While evidence is sparse, a recent review on *OXTR* DNAm by Moerkerke et al. (2021) ^26^ pointed towards a pattern of *hyper*methylation in adults with ASD ^27,28^, indicative of reduced *OXTR* expression. Contradictory, also patterns of *hypo*methylation were identified in children with ASD ^29,30^, suggesting differential ASD-related alterations in *OXTR* DNAm dependent on developmental stage. However, prior studies not only varied largely in terms of population-dependent characteristics (such as age), but also with regard to the investigated DNAm *OXTR* sites (e.g. intron versus exon CpG sites) as well as other design-related factors (e.g. adopted questionnaires/scales for assessing social/ASD-related traits) ^26^.

Despite these initial insights into the role of oxytocin hormonal variations and *OXTR* epigenetic modifications in ASD, research directly investigating the link between *OXTR* DNAm and hormonal oxytocin levels remains sparse. One study by Dadds et al. (2014) demonstrated that increased *OXTR* DNAm in peripheral blood was associated with lower plasma oxytocin levels in children/adolescents with oppositional-defiant/conduct disorder (4-to-16 year old) ^31^. However, another study in adults with psychotic disorders found no significant association between methylation of site −934 and plasma oxytocin ^32^. Likewise, a later study with female macaques also failed to identify any significant relationship between *OXTR* DNAm of distinct CpG sites and oxytocin levels in cerebrospinal fluid ^33^.

Finally, several lines of evidence also suggest a significant interplay between the oxytocinergic system and signaling of the hypothalamus-pituitary-adrenal (HPA) axis in stress regulation ^21^. For example, prior research in animals and humans provided indications of coupled releases of oxytocin and the HPA ‘stress-hormone’ cortisol, particularly in response to stressors, allowing oxytocin to inhibit stress-induced rises in cortisol ^34^. More research is needed however, to investigate these associations further, particularly in neuropsychiatric conditions such as ASD, in which elevated cortisol levels (see ^35^ for recent meta-analysis) and altered oxytocinergic signaling are apparent.

To fill these gaps, the present study aimed to examine possible alterations in peripheral hormonal levels of oxytocin and cortisol (assessed from saliva samples) as well as possible alterations in *OXTR* DNAm in a representative sample of school-aged children with and without ASD. Importantly, while endocrinological levels of e.g. cortisol are known to evolve during the course of the day ^36^ and in response to social encounters or stressful situations ^32,37^, this possibility of diurnal variations has been largely overlooked in prior research examining hormonal variations in oxytocin in ASD populations. It is however well-established that also oxytocin levels can covary depending on the impact of experimental (social) stimulations ^38^. The present study will therefore assess, for the first time, the interplay between oxytocin and cortisol hormonal levels in children with ASD, at two time points, either at ‘baseline’ in the morning or in the afternoon, after a (mildly stressful) social interaction session.

## Methods

This study was approved by the Ethical Committee for Biomedical Research at the University of Leuven, KU Leuven (S61358) in accordance with The Code of Ethics of the World Medical Association (Declaration of Helsinki). Written informed consent from the parents and assent from the child were obtained prior to the study.

### 1. Participants

Eighty children with a formal diagnosis of ASD, aged between 8-12 years, were recruited through the Leuven Autism Expertise Centre at the Leuven University Hospital between July 2019 and January 2021. Also, 40 age- and gender-matched typically developing (TD) peers were recruited.

Main inclusion criteria comprised a clinical diagnosis of ASD (only for children with ASD), premenstrual girls, intelligence quotient above 70 and native Dutch speaker. Main exclusion criteria comprised a history of any neurological disorder (stroke, concussion, epilepsy etc.), any significant physical disorder (liver, renal, cardiac pathology) or any neuropsychiatric diagnosis (only for TD children).

In addition, the Autism Diagnostic Observation Schedule (ADOS-2) ^39^ was acquired (**Table 1**). For all children, estimates of intelligence were acquired using four subtests of the Wechsler Intelligence Scale for Children, Fifth Edition, Dutch version ^40^ (**Table 1**).

**Table 1.** Participants’ characteristics.

|  | ASD group |  | TD group |  | Independent t-test |  |
| --- | --- | --- | --- | --- | --- | --- |
| | N | Mean ± SD | N | Mean ± SD | t-value | p-value |
| **Age** | 80 | 10.5 ± 1.3 | 40 | 10.3 ± 1.3 | 0.79 | 0.431 |
| **IQ** |  |  |  |  |  |  |
| Verbal IQ | 78 | 107.7 ± 15.2 | 40 | 117.3 ± 12.2 | -3.44 | < 0.001*** |
| Performance IQ | 79 | 102.3 ± 14.1 | 40 | 107.8 ± 12.2 | -2.09 | 0.039* |
| **Gender** |  |  |  |  |  |  |
| Girl | 16 (20%) |  | 8 (20%) |  |  |  |
| Boy | 64 (80%) |  | 32 (80%) |  |  |  |
| **Handedness** |  |  |  |  |  |  |
| Left | 10 (12%) |  | 6 (15%) |  |  |  |
| Right | 70 (88%) |  | 34 (85%) |  |  |  |
| **ADOS-2** |  |  |  |  |  |  |
| Social affect | 65 | 7.3 ± 3.7 | | / | | |
| Restricted and repetitive behavior | 65 | 1.9 ± 1.2 | | / | | |
| Total | 65 | 9.4 ± 4.1 | | / | | |
|  |  |  |  |  | MWU-test |  |
|  |  |  |  |  | Z | P-value |
| **ASD symptoms and behavioral problems** |  |  |  |  |  |  |
| Social responsiveness SRS-2 | 80 | 89.2 ± 21.3 | 40 | 21.9 ± 12.7 | 8.83 | <0.001*** |
| Repetitive/restrictive behavior RBS-R | 80 | 27.4 ± 15.7 | 40 | 2.5 ± 4.7 | 8.53 | <0.001*** |
| CBCL competences | 80 | 17.1 ± 4.5 | 40 | 22.9 ± 4.2 | -5.80 | <0.001*** |
| CBCL behavioral problems | 80 | 72.1 ± 23.9 | 40 | 21.9 ± 13.9 | 8.35 | <0.001*** |
|  |  |  |  |  | Independent t-test |  |
|  |  |  |  |  | t-value | p-value |
| **Biological samples** |  |  |  |  |  |  |
| OT levels |  |  |  |  |  |  |
| AM | 78 | 0.78 ± 0.65 | 39 | 1.07 ± 0.49 | -2.68 | 0.009** |
| PM | 77 | 0.96 ±0.69 | 40 | 1.02 ±0.49 | -0.49 | 0.629 |
| Cortisol levels |  |  |  |  |  |  |
| AM | 80 | -0.47 ±0.25 | 40 | -0.43±0.24 | -0.82 | 0.415 |
| PM | 78 | -0.91 ±0.24 | 40 | -0.95 ±0.26 | 0.84 | 0.400 |
| *OXTR* DNAm |  |  |  |  |  |  |
| CpG site -924 | 77 | 1.62 ±0.06 | 38 | 1.62 ±0.05 | 0.31 | 0.758 |
| CpG site -934 | 77 | 1.83 ±0.05 | 38 | 1.83 ±0.04 | -0.36 | 0.717 |

Data are shown as mean ± standard deviation. ASD autism spectrum disorder, TD typically developing, IQ Intelligence Quotient, ADOS-2 Autism Diagnostic Observation Schedule, SRS-2 Social Responsiveness Scale, RBS-R Repetitive Behavior Scale-Revised, CBCL Child Behavior Checklist, AM Anti Meridiem, PM Post Meridiem.
All biological samples were log-transformed.
\* $p < .05$
\*\* $p < .01$
\*\*\* $p < .001$

Children were also thoroughly characterized on autism symptom domains, using the parent-reported versions of the *Social Responsiveness Scale*, second edition (SRS-2) ^41,42^ and the *Repetitive Behavior Scale-Revised* (RBS-R) ^43^. Also, the parent-rated *Child Behavior Checklist* (CBCL) was obtained to assess behavioral problems and competences ^44^ (see **Supplementary Methods** for detailed descriptions of the adopted questionnaires).

As outlined in **Table 1**, groups were matched on age and sex, although verbal and performance intelligence quotient were overall higher in the TD group. As expected, children of the ASD group demonstrated significantly higher scores on the parent-rated SRS-2 and RBS-R, indicating more social impairments and more frequent expressions of restricted and repetitive behavior (*p* < .001, **Table 1**). The parent-rated CBCL also showed diagnosis-related effects, indicating more severe deficits in the ASD group, compared to the TD group (*p* < .05, **Table 1**). Note that the (biological) data collected for the current report were part of a larger clinical study including additional neurophysiological assessments (see also next section).

### 2. Oxytocin and cortisol salivary concentrations

For each child, salivary samples were acquired at two time points: (i) an AM sample, acquired at home, in the morning, within 30 minutes after awakening and before breakfast; and (ii) a PM sample, acquired in the afternoon, after completion of an experimental session performed at the Leuven University hospital. Importantly, the PM sample was collected within 30 minutes after finalizing an experimental test session, consisting of a semi-structured social interaction with an unknown experimenter, which could be experienced as moderately stressful, especially to the pediatric participants (see **Supplementary methods)**.

Salivary samples were collected using Salivette cotton swabs (Sarstedt AG & Co.). For the analysis of salivary **oxytocin levels**, the commercial enzyme immunoassay oxytocin ELISA kit of Enzo Life Sciences, Inc. was used, similar to prior studies ^45–47^. All sample extraction and concentration procedures were conducted in accordance with manufacturer’s instructions. Measurements were performed on undiluted samples (100 μl), and sample concentrations were calculated according to plate-specific standard curves.

Analysis of the **cortisol levels** was performed using the commercial enzyme immunoassay Cortisol ELISA kit of Salimetrics, Europe. Measurements were performed on undiluted samples (25 μl), and sample concentrations were calculated according to plate-specific standard curves. More detailed information regarding the salivary collection procedures and analyses is provided in **Supplementary methods**.

### 3. DNA methylation of the oxytocin receptor gene

Additional salivary samples were obtained via the Oragene DNA sample collection kit (DNA Genotek Inc., Canada) to address epigenetic variations in the level of methylation (DNAm) at two CpG sites (−934 and -924) of *OXTR* (hg19, chr3:8,810,729-8,810,845).

After data collection, 200 ng DNA was extracted from the samples and bisulfite converted following the manufacturer’s protocol (EZ-96 DNAm Kit, Zymo Research, Irvine, CA, USA). Bisulfite converted DNA was stored at −80 °C until further analysis. Next, the levels of methylation at two CpG sites (i.e., -934 and -924) of *OXTR* (hg19, chr3:8,810,729-8,810,845) were determined using Pyrosequencer (Qiagen, Hilden, Germany) and analyzed using Pyromark Q96 software. Laboratory procedures and analyses were conducted in accordance with manufacturer’s protocols and software settings (e.g. for determining unreliable samples) ^44^. Protocols for the PCR amplification and Pyrosequencing analysis were adapted from Krol et al. (2019) ^48^. More information regarding the adopted PCR primers can be found in **Supplementary methods**.

### 4. Data handling and statistical procedures

Prior to analysis, hormonal and DNAm data were log-transformed (log10) to deal with skewed data. Next, diagnosis-related differences were assessed by subjecting oxytocin and cortisol hormonal data to repeated-measure analyses-of-variance (ANOVA) with the between-subject factor group (ASD, TD) and the within-subject factor time point (AM, PM) (see **Supplementary table 1-2** for full ANOVA models).

*OXTR* DNAm data were subjected to independent sample t-tests to examine diagnosis-related differences in DNAm at CpG site -924 and -934.

Pearson correlation analyses were performed to assess associations between the two hormonal systems in children with or without ASD and between hormonal levels and *OXTR* DNAm. All statistical analyses were executed with SPSS (version 28.0, IBM).

## Results

### Diagnosis-related differences

#### Oxytocin levels

A repeated-measure ANOVA revealed no main effects of group (*F*(1,113) = 2.45; *p* = .120) or time-point (*F*(1,113) = 0.87; *p* = .354). A significant group x time point interaction (*F*(1,113) = 4.23; *p* = .042) was noted, indicating significantly lower morning oxytocin levels in ASD, compared to TD children (AM sample, independent-samples *t*(115) = - 2.68; *p* = .009, **Figure 1A and Table 1**) but no differential afternoon oxytocin levels in ASD, compared to TD children (PM sample *t*(115) = -0.49; *p* = .629, **Figure 1A and Table 1**). Notably, while oxytocin levels remained overall constant from AM to PM in the TD group (dependent-samples *t*(38) = -0.67; *p* = .507), the ASD group displayed a steep and significant increase in oxytocin levels from AM to PM (*t*(75) = 2.61; *p* =.011) (**Figure 1A**).

**Figure 1.**
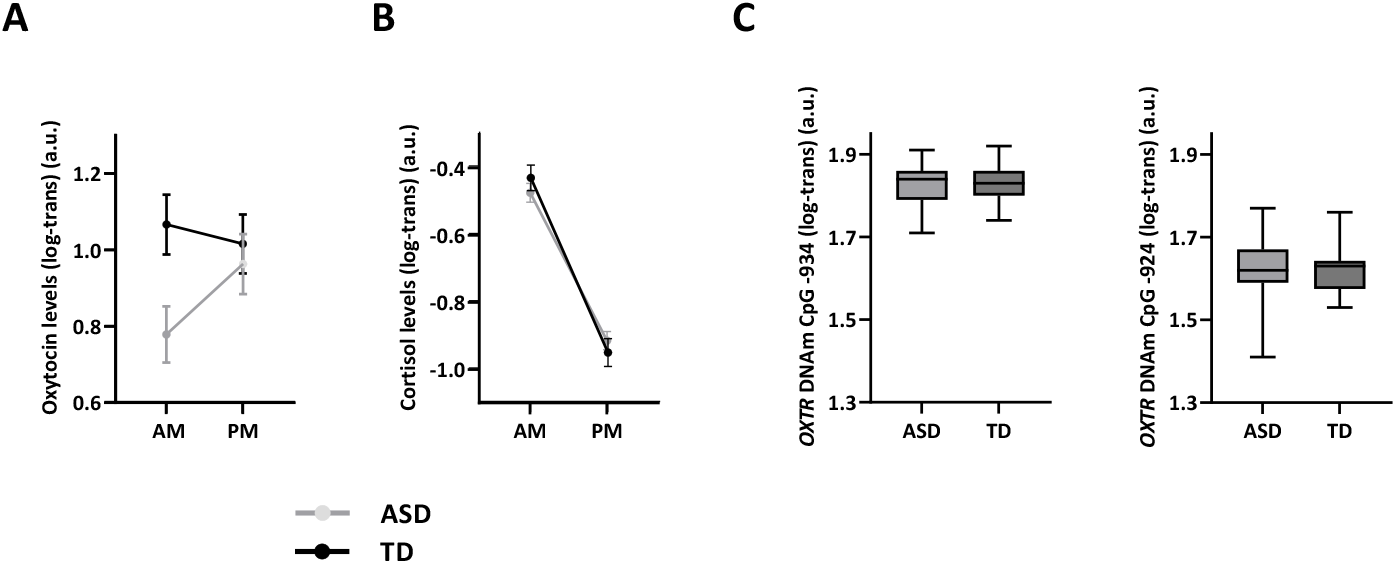
Diagnosis-related differences. Salivary oxytocin (**panel A**) and cortisol hormonal levels (**panel B**), as well as OXTR gene methylation data (**panel C**) are visualized separately for children with ASD and typically developing children (TD). Hormonal concentrations are visualized separately for the morning sample (AM), and the afternoon sample (PM), acquired after a mildly stress-inducing social interaction session. Methylation frequency of the OXTR gene is visualized separately for CpG site -934 and CpG site -924. **Panel A**. Oxytocin levels were significantly higher in the TD group, compared to the ASD group, for the morning AM sample, not for the PM sample collected after the social interaction session. While oxytocin levels remained overall constant across AM and PM measurements in the TD group, the ASD group displayed a steep increase in oxytocin levels from the AM measurement to the PM measurement. **Panel B**. Cortisol levels were not significantly different between the ASD and the TD group. Both groups displayed a similar cortisol peak in the morning and a similar level of cortisol at the PM measurement, after finalizing the social interaction session. **Panel C**. *OXTR* DNAm at CpG sites -934 and -924 were not significantly different between the ASD group and the TD group.

#### Cortisol levels

A repeated-measure ANOVA revealed no significant main effect of group (*F*(1,116) = .002; *p* = .965) nor group x time point interaction (*F*(1,116) = 1.86; *p* = .175) (**Figure 1B and Table 1**). A significant main effect of time point was evident however (*F*(1,116) = 286.83; *p* < .001), indicating that in both groups, a steep decline in cortisol levels was evident from AM to PM. Accordingly, it appears that - as a group - children with or without ASD display a similar cortisol peak in the morning and a similar pattern of cortisol diminishment during the course of the day.

### *OXTR* DNAm

No significant diagnosis-related differences in *OXTR* DNAm were evident between the ASD and TD group, either at CpG site -934 (*t*(113) = -0.33; *p* = .745) or at CpG site -924 (*t*(113) = 0.44; *p* = .659) (**Figure 1C**).

### Association between oxytocin and cortisol hormonal levels

In the TD group, a tight interaction between oxytocin and cortisol hormonal systems was evident, indicating that high morning oxytocin levels (AM sample) were predictive of lower ‘stress-induced’ cortisol levels (PM sample) (Pearson *r* = -.35; *p* = .027) (**Figure 2A**). Higher oxytocin levels in the afternoon, i.e. after the (mildly stressful) social interaction session, were also significantly associated with reduced cortisol levels in the afternoon (PM sample) (*r* = - .34; *p* = .032) (**Figure 2B**).

**Figure 2.**
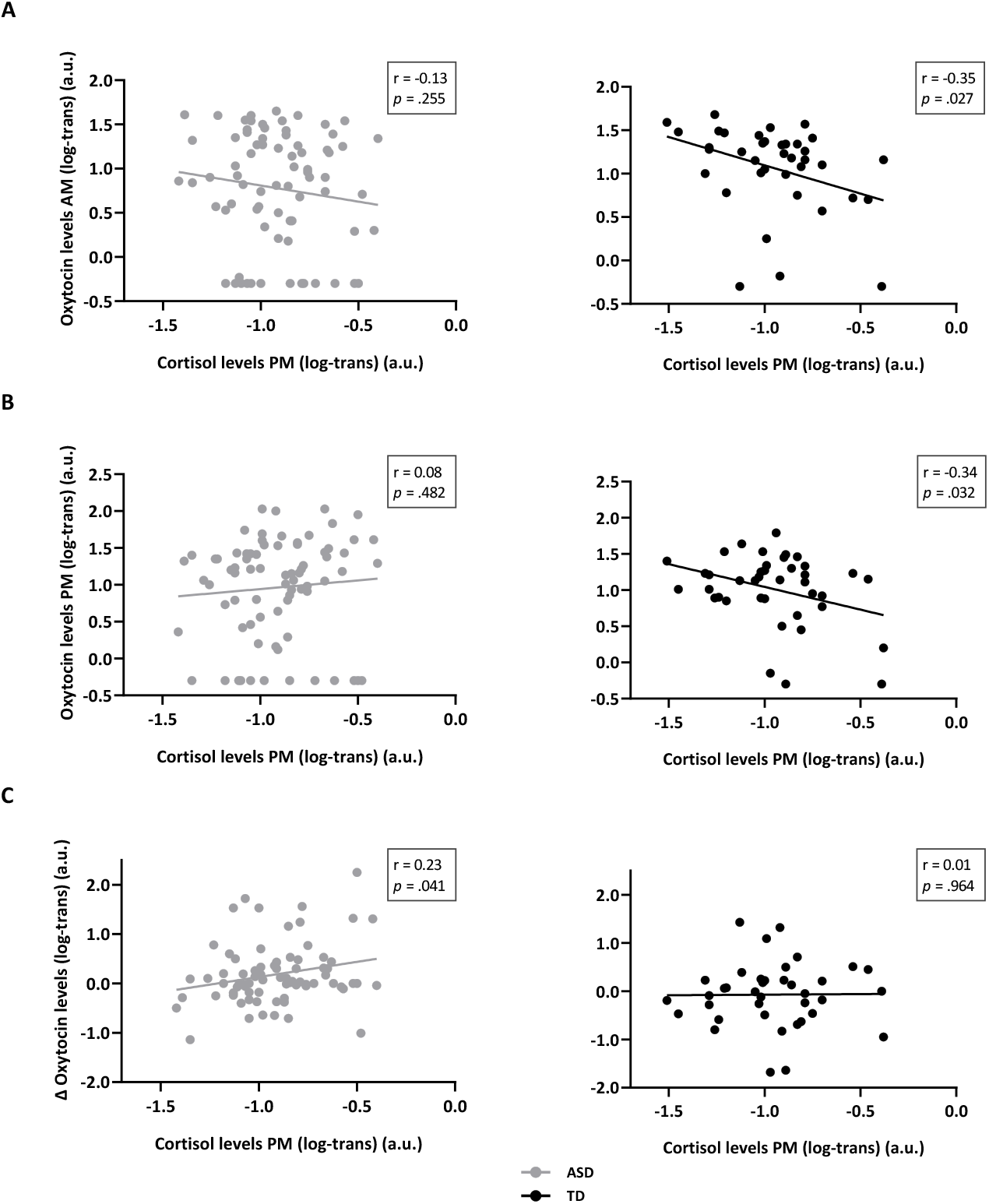
Association between oxytocin and cortisol hormonal levels. Relationships between oxytocin and cortisol hormonal levels are visualized separately for children with ASD and typically developing children (TD). **Panel A**. In the TD group, oxytocin levels at AM were significantly associated with cortisol levels at PM, indicating that high morning oxytocin was predictive of low ‘stress-induced’ PM cortisol levels. Relationships were only significantly evident in the TD group, not in the ASD group. **Panel B**. Oxytocin levels at PM were also significantly associated with cortisol levels at PM, indicating that higher afternoon oxytocin levels were associated with lower ‘stress-induced’ cortisol levels. Also here, the relationship was only evident in the TD group, not in the ASD group. **Panel C**. Finally, in the ASD group, the observed marked increases in oxytocin levels during the day (from the AM to the PM assessment) were significantly associated with higher afternoon ‘stress-induced’ cortisol levels.

Strikingly, these associations were only evident in the TD group, not in the ASD group, suggesting a lack of coupling of these hormonal systems in children with ASD (all, *r* < .13; *p* > .05). **Supplementary table 3** reports a full overview of the assessed associations between oxytocin and cortisol levels, separately for each group, including a direct comparison of correlation values between ASD and control groups.

Notably, considering the marked AM to PM increase in oxytocin levels seen in the ASD group, we additionally explored whether these AM to PM changes in oxytocin levels were potentially related to cortisol levels. It was revealed that indeed, only in the ASD group (*r* = .23; *p* = .041), not in the TD group (*r* = .01; *p* = .964), higher cortisol levels at PM were associated with a significant rise in oxytocin levels during the day (from AM to PM) (**Figure 2C**).

### Associations between hormonal levels and OXTR DNAm

#### CpG site -924

Only in the TD group (*r* = -.41; *p* = .011), not in the ASD group (*r* = .05; *p* = .682), a significant association between *OXTR* DNAm of site -924 and PM cortisol levels was evident, indicating that children with lower methylation frequency of CpG site -924 (higher *OXTR* expression) displayed higher afternoon cortisol levels (**Figure 3**). *OXTR* DNAm of site -924 was not significantly associated with AM cortisol levels or AM/PM oxytocin levels either in children with or without ASD (all, *r* < .07; *p* > .05; see **Supplementary table 3** for a full overview).

**Figure 3.**
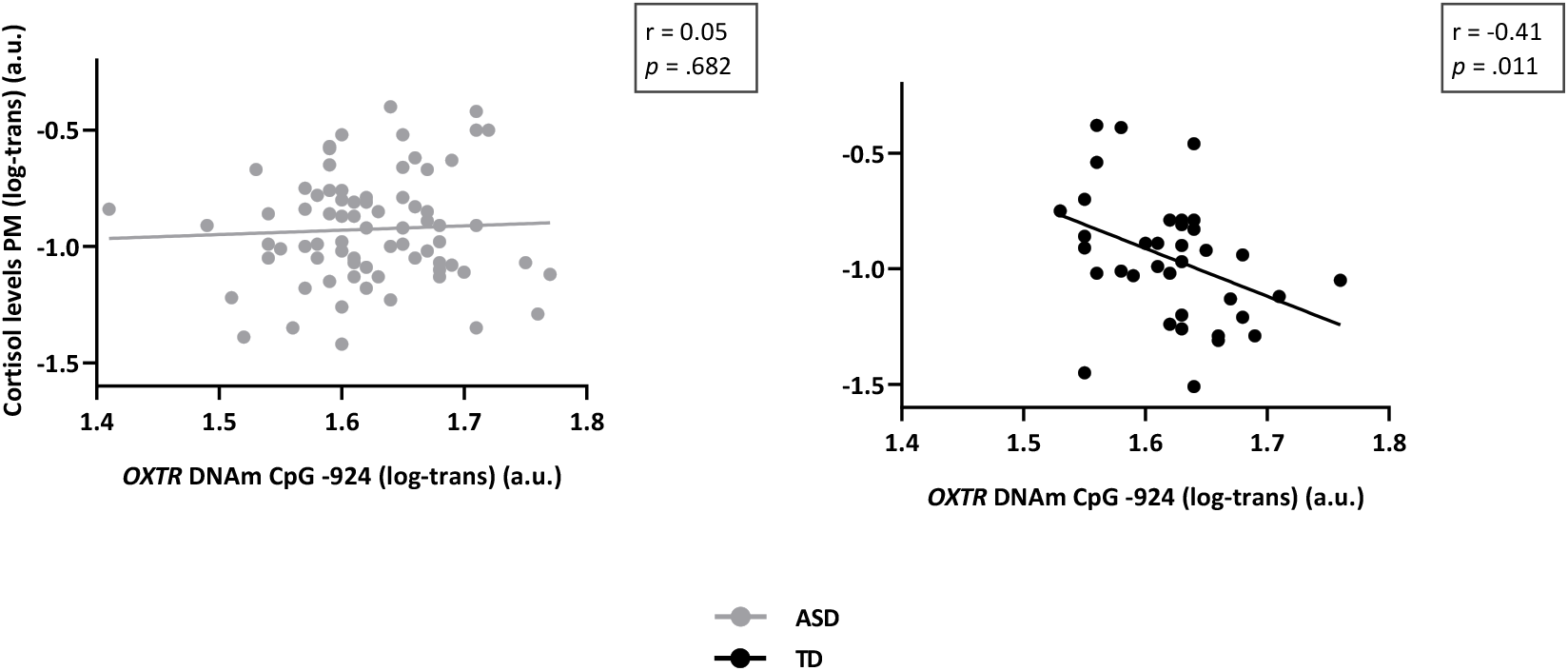
Associations between hormonal levels and OXTR DNAm. Associations between hormonal levels and methylation of the *OXTR*, visualized separately for children with and without ASD. In the TD group, not in the ASD group, a significant negative association was evident between *OXTR* methylation of CpG site -924 and cortisol levels, indicating that children with lower methylation frequency (higher *OXTR* expression) displayed higher afternoon cortisol levels.

#### CpG site -934

Methylation of CpG site -934 was not significantly associated with AM or PM oxytocin or cortisol levels, either in children with or without ASD (all, *r* < .08; *p* > .05, see **Supplementary table 3**).

## Discussion

In the current study, we investigated alterations in oxytocin and cortisol hormonal systems as well as *OXTR* DNAm in school-aged children with ASD, compared to a group of age and gender matched TD children.

Only in terms of ‘baseline’ oxytocin, children with ASD displayed reduced levels of peripheral oxytocin assessed in the morning, not in terms of afternoon oxytocin levels, assessed after induction of a mildly stressful social interaction session. These findings therefore provide indications that only *‘trait-dependent’* oxytocin levels, but not stress-reactive *‘state-dependent’* levels of oxytocin are altered in ASD. Further, while overall cortisol levels were similar in children with and without ASD, differential couplings between the oxytocin and cortisol hormonal system were evident, indicating that only in the TD group, not in the ASD group, higher oxytocin levels were predictive of dampened afternoon ‘stress-induced’ cortisol levels. Finally, DNAm at two distinct CpG sites of *OXTR* were not altered in children with ASD, although note that only in the TD group, methylation frequency was significantly associated with hormonal reactivity of the cortisol system.

Diagnosis-related differences in oxytocin hormonal levels were evident in children with ASD, albeit exclusively in terms of ‘trait-dependent’ morning oxytocin, not in terms of ‘state-dependent’ afternoon oxytocin levels, measured after an experimental social interaction session. Prior meta-analytic analyses examining altered oxytocin levels in ASD yielded a mixed pattern of results, predominantly depending on the age of the included participants ^12^. While it is difficult to directly compare studies, the current data provide important indications that also the time of the day as well as the particular experimental and social context (with or without experimental manipulation) are important factors to consider for explaining possible variations in oxytocin levels.

Closer inspection of the recent meta-analysis of Moerkerke et al. (2021) ^12^ showed that only in 11 out of 18 studies the timepoint or time range of sampling was reported. Of these studies, six reported to have acquired ‘morning’ oxytocin samples (AM) ^13,15,17,20,21,49^, and four of these six reported similar group-related differences as in our study, indicating lower morning oxytocin levels in ASD compared to the TD group ^13,15,21,49^. In accordance with our study, the only two studies who indicated to have sampled oxytocin in the afternoon (PM), reported no significant group differences ^18,19^. Further, in the remaining studies who reported sampling timings, it appeared that acquisition timing was not standardized across individuals (i.e., ranging from morning to afternoon), therefore rendering direct comparisons with our current results was difficult ^22,50,51^. As the time point and context of sampling may have an important impact on observed levels of oxytocin and correlations with other outcome measures, future studies are urged to report this important design-specific information in more detail.

With regard to cortisol levels, in the current study, no differences in morning or afternoon cortisol levels were identified in children with or without ASD, indicating no overall alterations in HPA axis reactivity, as demonstrated before (^21^, although see ^52^). Notably, depending on diagnosis, differential couplings between the oxytocinergic system and the HPA axis (as indexed with cortisol) were evident, indicating that only in the TD children, not in children with ASD, heightened (morning) oxytocin levels were predictive of dampened ‘state-dependent’ release of cortisol. Particularly, this negative coupling was strongest between morning oxytocin and afternoon cortisol levels measured after the mildly stress-inducing social interaction session. These results therefore suggest that higher ‘trait-dependent’ levels of morning oxytocin, seen in TD children, may constitute an important *protective* mechanism to buffer HPA axis activity upon (social) stressors encountered during the day. In children with ASD, on the other hand, this protective oxytocinergic HPA-stress attenuation mechanism may be dysfunctional or absent, considering the overall diminished levels of morning oxytocin in the children with ASD, and the apparent lack of coupling between oxytocin and cortisol systems in this group.

Overall, our observations of oxytocinergic dampening of HPA - cortisol responses are in line with prior studies showing acute, dampening effects of intranasal oxytocin administration on cortisol reactivity to laboratory tasks ^53,54^. Interestingly, meta-analytic evidence showed that the attenuating effects were most pronounced in response to challenging laboratory tasks that produced a robust stimulation of the HPA axis, as well as in clinical populations with ASD or schizophrenia, compared to neurotypicals ^53^. This latter observation is important, as the authors interpreted this finding to reflect an increased sensitivity to the exogenously administered oxytocin among those with a clinical diagnosis, presumably because they may present lower baseline oxytocin levels. Albeit indirectly, our current observations corroborate this interpretation by showing that morning levels of salivary oxytocin are indeed significantly diminished in children with ASD. Exogenously administered oxytocin may therefore be particularly effective in ASD, i.e., for restoring the otherwise dysfunctional oxytocinergic HPA stress attenuation.

Notably, while diminished levels of oxytocin were initially noted in children with ASD, a significant increase was observed from AM to PM, reaching a similar level of concentration as noted in TD children. In TD children, levels remained overall stable throughout the day. While speculative, the steep increase in oxytocin release in ‘state-dependent’ afternoon oxytocin, compared to ‘trait-dependent’ morning oxytocin, can be hypothesized to reflect a compensatory mechanism of *reactive* oxytocin release upon the mildly stressful social interaction session. Indeed, while absolute oxytocin and cortisol levels were not significantly associated in children with ASD, exploratory analyses confirmed that, in the ASD group, higher state-dependent, afternoon cortisol levels were paralleled by a steeper morning to afternoon rise in oxytocin levels, likely reflective of a coupled co-release of cortisol and oxytocin upon stress-inducing tasks, as demonstrated before in both animals ^55^ and humans ^34^. In this view, the parallel release of oxytocin upon HPA activation may be considered a *reactive* stress-regulatory mechanism, which - over time - may facilitate a dampening and further reduction of the cortisol-induced stress response. As highlighted by Brown et al. (2016), positive associations between oxytocin and cortisol levels upon acute stress induction are thought to be reflective of an initial co-release, for subsequently dampening stress responses and facilitating coping behaviors ^34^. These and our findings therefore highlight the importance of studying the interaction among oxytocinergic and HPA stress systems from a dynamic, time-varying perspective, rather than studying static levels of a single marker.

To sum up, in children with ASD, the *positive* association between the steep rise in oxytocin levels and stress-induced HPA-cortisol levels may be indicative of a *reactive* stress-regulatory mechanism. In TD children, on the other hand, the identified *negative* association between trait (morning) levels of oxytocin and stress-induced HPA-cortisol levels are likely reflective of a *protective* oxytocinergic stress-regulatory mechanism.

Aside hormonal characterizations, also variations in DNAm frequencies of distinct CpG sites of *OXTR* were assessed, but no significant group differences were identified. These results accord to observations of a recent study by Siu et al. (2021), neither showing significant group differences in the level of methylation of CpG sites -934 and -924 in children and adolescents with and without ASD ^30^. Despite some variation, a recent review pointed towards differential patterns of hypo-versus hyper-methylation, respectively in children and adults with ASD, although the number of included studies was limited, and overall sample sizes were small to modest ^26^. Also, considerable design-related variations were noted by the authors, *e*.*g*. in terms of included CpG sites, rendering it difficult to draw conclusive interpretations regarding ASD diagnosis-related alterations in *OXTR* DNAm based on the existing evidence.

In terms of relationships between hormonal levels and *OXTR* DNAm, only the TD group displayed a significant negative association between *OXTR* DNAm (at CpG site -924) and state-dependent (afternoon) cortisol levels, indicating that lower methylation frequencies were associated with higher cortisol levels. This finding accords to a prior study identifying a similar negative relationship between stress-induced cortisol and *OXTR* DNAm frequency in individuals with social anxiety disorder (SAD) ^56^. In the latter study, the pattern of *OXTR* hypomethylation associated with SAD was interpreted to be reflective of a compensatory mechanism, i.e., facilitating an upregulation of oxytocin receptors to cope with heightened HPA axis activity in SAD. More research is needed however, to corroborate this interpretation and pattern of results in future studies.

While the current study provides important new insights into the oxytocinergic system of children with ASD and its interactions with HPA stress reactivity, the following limitations and recommendations are noted. First, as indicated, the included children reflected a rather homogenous group of high-functioning children within a tight pre-pubertal age range, rendering generalizability of the identified effects to more heterogeneous and/or younger/older children or adolescents uncertain. Also, while the included number of boys and girls in our sample reflected the well-documented four-to-one male bias in ASD prevalence ^57^, future research is warranted to examine the observed effects also in larger samples of girls with ASD, especially given prior reports of gender-related differences in oxytocinergic function ^58^. Further, while the inclusion of multiple salivary sampling time points (both in the morning and afternoon) was a strong asset for examining dynamic trait (morning) versus state-dependent (after experimental manipulation) hormonal levels, it is noted that any future design would benefit from amplifying the sampling frequency even further, allowing an even more fine-grained sampling of dynamic changes in hormonal systems interactions ^34^.

To conclude, this study provides new insights into altered oxytocinergic signaling in children with ASD and its interactions with the cortisol HPA stress system. Results provide important indications that only ‘trait-dependent’ oxytocin levels, but not (social) stress-induced ‘state-dependent’ levels of oxytocin are altered in ASD. These observations suggest that TD children without ASD may rely more on a *protective* stress-regulatory mechanism, involving high circulating trait levels of oxytocin for buffering stress reactivity, whereas children with ASD may rely more heavily on a *reactive* stress regulatory mechanism, involving more pronounced stress-induced, state-dependent releases of oxytocin upon HPA - cortisol activation, i.e., for reactively dampening stress responses and facilitating coping behaviors. While more work is needed, the identified results may aid in establishing relevant biomarkers for diagnostic and/or treatment evaluation purposes, as well as for the development of new (personalized) treatment approaches for targeting the oxytocinergic system in ASD.

## Data Availability

All data produced in the present study are available upon reasonable request to the authors

## Acknowledgments

This research was supported by internal funding of the KU Leuven (ELG-D2857-C14/17/102), a Doctor Gustave Delport fund of the King Baudouin Foundation, a Branco Weiss fellowship of the Society in Science - ETH Zurich, and an EOS Excellence of Science grant (G0E8718N, HUMVISCAT) granted to KA and BB. ME is supported by an FWO aspirant fundamental fellowship [11N1222N]. JP is supported by an FWO junior postdoctoral fellowship [1257621N].

The funding sources had no further role in study design; in the collection, analysis and interpretation of data; in the writing of the report; and in the decision to submit the paper for publication.

