## Supplementary material for "Endogenous oxytocin levels in children with autism: Associations with cortisol levels and oxytocin receptor gene methylation"

#### Title

### Supplementary methods

#### Questionnaires

Children of the ASD and the TD group were thoroughly characterized using questionnaires assessing autism symptom domains, and behavioral problems and competences. Questionnaire scores are listed in **Table 1**, separately for the ASD and TD group.

Difficulties in the social domain were assessed using the parent-report **Social Responsiveness Scale**, second edition (SRS-2), comprising 65 items scored on a four-point Likert-scale, with higher scores indicating a greater deficit. Repetitive and restrictive behavior were assessed using the parent-report **Repetitive Behavior Scale** - Revised (RBS-R), comprising 43 items scored on a four-point Likert-scale, with higher scores indicating greater deficits.

Overall behavioral problems and competences were measured using the parent-rated **Child Behavior Checklist** (CBCL). The competence subscale consists of 20 items, scored on a four-point Likert-scale, with a higher score relating to better school/social competences and participation in activities in leisure time. The behavioral problems subscale consists of 120-items, scored on a three-point Likert-scale, with a  higher score indicating a higher incidence and severity of behavioral problems .

All questionnaire data were acquired and stored using a secure web-based application (MyNexuzhealth) and Research Electronic Data Capture software (REDCap).

**Hormonal samples**

***Collection.*** Participants were asked to chew the cotton swab for approximately 60 seconds until it was saturated with saliva. Before collection, participants were asked to subject to oral resting (no eating, chewing gum, or drinking) for approximately 30 minutes. After collection, swabs were placed in a sterile conical tube and placed in a −20°C freezer for storage up until centrifugation.

***Sampling timings.*** AM samples were on average collected at 08:07(range 06:40 – 11:40) for the ASD group, and at 07:52 (range 06:00- 09:15) for the TD group (Watson-Williams test (circular statistics): $F$ = 2.34 ; $p$ = .129). Afternoon samples were on average collected at 15:34 (range 11:45– 19:05) for the ASD group, and at 14:50 (range 11:37– 18:40) for the TD group (Watson-Williams test (circular statistics): $F$ = 3.33; $p$ = .071).

***Experimental session.*** The PM samples were collected within 30 minutes after finalizing a mildly stressful social interaction session. It consisted of a semi-structured social interaction with an unknown experimenter, involving eye contact and a short conversation about the child's interests and everyday life experiences. Furthermore, a 45-minute MRI protocol was administered (involving a series of structural and functional scans) as well as a 45-minute EEG and physiology assessment (involving being touched by a stranger on the head, the face and the breast to position the EEG cap and the ECG sensors).

***Available oxytocin samples.*** Home collected AM samples were available for all children (80 ASD, 40 TD). PM samples were not collected for two children of the ASD group. Further, due to COVID-19 restrictions during data collections, for 10 ASD children, PM samples were collected at home, thus without undergoing the stressful lab experience.

Note however, that the main patterns of results were not qualitatively impacted when performing the analyses with or without these participants.

Oxytocin levels could not be determined for three subjects (2 ASD, 1 TD) in the morning (AM), and for three subjects (3 ASD) in the afternoon (PM), due to insufficient saliva fluid. Salivary samples with only 50 µl of sample material available were diluted to 100 µl and sample concentrations were multiplied by 2 (4 AM and 1PM sample of the ASD group). Further, oxytocin level data of 5 participants (2 AM and 3 PM samples of the ASD group) were identified as extreme outliers and were recoded to the mean (across groups) (three inter-quartile ranges (Q3-Q1) below or above the first (Q1), respectively third (Q3) quartile). Finally, for participants displaying concentrations below the detection limit, concentrations were set to the lowest detected value across samples (i.e. 14 AM and 12 PM samples of the ASD group; 2 AM and 2 PM samples of the TD group). Note that also here, the main patterns of results were not qualitatively impacted when performing the analyses with or without these participants.

***Available cortisol samples.*** For the cortisol data, 7 participants were identified as extreme outliers (2 AM samples and 1 PM sample of the ASD group; 3 AM samples and 1 PM sample of the TD group) and were recoded to the mean (across groups).

**DNAm samples**

***DNAm sampling timings.*** Oragene DNA saliva samples were on average collected at 15:37 (range 11:50 – 19:07) for the ASD group, and at 14:47 (range 10:25 – 18:40) for the TD group (Watson-Williams test (circular statistics): $F = 4.15$; $p = .044$).

***Available DNAm samples.*** Salivary samples for characterization of *OXTR* DNAm were obtained from all participants, except for one child of the ASD group. *OXTR* DNAm data could not be obtained for three additional participants of the ASD group and two participants of the TD group due to insufficient sample quality. Additionally, data of one participant of the ASD group was identified as an outlier and was recorded to the variable mean (for both CpG sites).

Salivary collections constitute a fairly simple and stress-free method that is easily applicable in vulnerable (pediatric) populations. Furthermore, recent evidence indicates that it is as reliable and suitable for DNAm analyses as methylation patterns detected in other tissues such as brain and blood [1].

**_OXTR_ DNAm primers**

The following PCR primers were adopted to amplify the DNA fragment of interest at the CpG sites -934 and -924: [OXTR Forward: TTG AGT TTT GGA TTT AGA TAA TTA AGG ATT; OXTR Reverse: /5Biosg/AC TTA ACA TCA CAT TAA ATA CAA CC]. The PCR conditions during the amplification were as follows: step 1: (95°C/15 min)/1 cycle; step 2: (94°C/30 s, 58°C/30 s, 72°C/30 s)/50 cycles; step 3: (72°C/10 min)/1 cycle; and step 4: 4°C hold. Also the following sequencing primer was used to read the DNA sequence (OXTR Sequencing: AGA AGT TAT TTT ATA ATT TT).

#### Supplementary table 1.

*Repeated measure ANOVA model of the oxytocin hormonal levels (log-transformed)*

| | Sum of squares | df | Mean square | F | p-value |
|---|---|---|---|---|---|
| Intercept | 185.934 | 1 | 185.934 | 331.826 | <.001** |
| Group | 1.377 | 1 | 1.377 | 2.458 | .120 |
| Error | 63.318 | 113 | 0.560 | | |
| Time | 0.174 | 1 | 0.174 | 0.865 | .354 |
| Time*Group | 0.852 | 1 | 0.852 | 4.234 | .042* |
| Error (time) | 22.728 | 113 | 0.201 | | |

*  *p*<.05

** *p*<.001

#### Supplementary table 2.

*Repeated measure ANOVA model of the cortisol hormonal levels (log-transformed)*

| | Sum of squares | df | Mean square | F | p-value |
|---|---|---|---|---|---|
| Intercept | 101.804 | 1 | 101.804 | 1293.960 | <.001** |
| Group | 0.000 | 1 | 0.000 | 0.002 | .965 |
| Error | 9.126 | 116 | 0.079 | | |
| Time | 12.259 | 1 | 12.259 | 286.829 | <.001** |
| Time*Group | 0.080 | 1 | 0.080 | 1.863 | .175 |
| Error (time) | 4.958 | 116 | 0.043 | | |

** *p*<.001

#### Supplementary table 3.

*Correlation analyses assessing relationships between oxytocin hormonal levels and biological samples (cortisol, OXTR DNAm) and behavior assessments of autism symptoms and attachment. Correlations are reported separately for the AM and PM sample, and for each diagnostic group (ASD, TD).*

| | ASD group | | | TD group | | | Comparison correlation coefficients | |
|---|---|---|---|---|---|---|---|---|
| | Correlation coefficient (r) | *p*-value | N | Correlation coefficient (r) | *p*-value | N | Z-value | *p*-value |
| **Oxytocine levels AM** | | | | | | | | |
| **Biological samples** | | | | | | | | |
| Cortisol level AM | 0.04 | .711 | 78 | -0.29 | *.075* | 39 | 1.67 | .094 |
| Cortisol level PM | -0.13 | .255 | 76 | -0.35 | **.027*** | 39 | 1.16 | .246 |
| DNAm -924 | -0.07 | .560 | 75 | -0.01 | .952 | 37 | | |
| DNAm -934 | -0.08 | .494 | 75 | -0.07 | .701 | 37 | | |
| **Oxytocine levels PM** | | | | | | | | |
| **Biological samples** | | | | | | | | |
| Cortisol level AM | 0.10 | .380 | 77 | -0.30 | *.057* | 40 | 2.07 | **.039*** |
| Cortisol level PM | 0.08 | .482 | 77 | -0.34 | **.032*** | 40 | 2.16 | **.031*** |
| DNAm -924 | 0.01 | .971 | 74 | 0.29 | *.077* | 38 | -1.43 | .154 |
| DNAm -934 | 0.07 | .585 | 74 | 0.01 | .955 | 38 | | |

Pearson correlation analyses

ASD autism spectrum disorder, TD typically developing, AM Anti Meridiem, PM Post Meridiem, DNAm DNA methylation.

All biological samples data was log-transformed.

* $p<.05$
